# Next generation plasma proteome profiling of COVID-19 patients with mild to moderate symptoms

**DOI:** 10.1101/2021.06.15.21258940

**Authors:** Wen Zhong, Ozlem Altay, Muhammad Arif, Fredrik Edfors, Adil Mardinoglu, Mathias Uhlén, Linn Fagerberg

## Abstract

COVID-19 has caused millions of deaths globally, yet the cellular mechanisms underlying the various effects of the disease remain poorly understood. Recently, a new analytical platform for comprehensive analysis of plasma protein profiles using proximity extension assays combined with next generation sequencing has been developed. Here, we describe the analysis of the plasma profiles of COVID-19 patients with mild and moderate symptoms by comparing the protein levels in newly diagnosed patients with the plasma profiles in the same individuals after recovery 14 days later. The study has identified more than 200 proteins that are significantly elevated at infection and many of these are related to cytokine response and other immune-related functions. In addition, several other proteins are shown to be elevated, including SCARB2, a host cell receptor protein involved in virus entry. A comparison with the plasma protein response in patients with severe symptoms shows a highly similar pattern, but with some interesting differences. In conclusion, the results will facilitate further studies to understand the molecular mechanism of the immune-related response of the SARS-CoV-2 virus.

## INTRODUCTION

Corona virus disease 2019 (COVID-19) caused by the severe acute respiratory syndrome coronavirus-2 (SARS-CoV-2) is a highly contagious disease. Patients infected with COVID-19 suffers from a large variation of symptoms caused by the host immune response, including substantial respiratory problems, acute coronary syndromes and metabolic dysfunction (Del Valle *et al*, 2020; Gupta *et al*, 2020; Lucas *et al*, 2020; Mathew *et al*, 2020). The mechanisms behind the disease and why some remain asymptotic carriers while other patients experience severe disease with fatal outcome are poorly understood (Williamson *et al*, 2020), however, many recent studies have suggested that cytokine storms and immunosuppression is highly associated with progression of the disease (Hou *et al*, 2020; Huang *et al*, 2020; Mehta *et al*, 2020; Wu & McGoogan, 2020).

An important effort to understand the biology of the host-virus response is to move towards comprehensive proteome profiling of host proteins in blood in response to viral infections, not only to understand the basis for disease, but also to facilitate precision medicine efforts aimed at stratification and monitoring of patients before and during therapeutic interventions (Gummesson *et al*, 2021; Zhong *et al*, 2021; Zhong *et al*, 2020). The objective is thus to probe the circulating plasma proteome of individuals with sensitive and specific assays that can allow massive sample throughput. However, progress has been hampered by the challenge to allow the quantification of thousands of proteins across more than a billion range in concentrations, starting with minute sample volumes (Zhong *et al*., 2021).

We have previously reported the stable and unique plasma proteome profiles in healthy individuals (Tebani *et al*, 2020; Zhong *et al*., 2020), based on the Proximity Extension Assay (PEA) method (Assarsson *et al*, 2014). Recently, we have also shown that this can be extended for simultaneous analysis of many more targets by the introduction of massive parallel sequencing, here referred to as PEA-NGS, without sacrifice on accuracy or sensitivity (Zhong *et al*., 2021). This new approach for “next generation plasma profiling” allows for simultaneous analysis of close to 1500 protein targets from small volumes of samples and facilitates sensitive multiplex assays to be coupled with low cross-reactivity and minimal off-target events, as exemplified by the analysis of type 2 diabetes patients (Zhong *et al*., 2021). Recently, Patel et al (Patel *et al*, 2021) and Filbin et al (Filbin *et al*, 2021) described using the PEA analytical platform the plasma protein profiling of COVID-19 patients with a main focus on individuals with severe symptoms. Here, we have used the PEA-NGS analysis to investigate the plasma proteome profile of COVID-19 patients with mild to moderate symptoms to allow comprehensive comparisons of protein responses as a result of infection.

## RESULTS

### The study cohorts

The analysis includes a cohort of 50 individuals with an ongoing COVID-19 infection and the plasma profiles in these individuals are compared with a healthy control population. Patients were recruited with a positive PCR test for SARS-CoV-2 and blood samples taken for analysis within 24 hours of confirmation of a COVID-19 infection (day-0) and after 14 days (day-14). We have previously reported the individual plasma proteome variation in a healthy cohort with individuals between 50 - 65 years as part of the Swedish SCAPIS SciLifeLab Wellness profiling program (S3WP) (Tebani *et al*., 2020; Zhong *et al*., 2021; Zhong *et al*., 2020), and this cohort was here used to allow for a comparison with a healthy control population. The study design is shown in **Fig 1A**. The COVID-19 cohort consisted of individuals with a wide range in age (19 to 66) (**Fig 1B**) with an average of 38 years and with an average body mass index (BMI) of 27 (18.8 to 37.8) (**Fig 1C**). The number of days with symptoms after positive PCR-test is shown in **Fig 1D**, with an average of 7.5 days. All individuals suffered from mild to moderate symptoms due to COVID-19 and a summary of the respective symptoms as well as the measured oxygen saturation (SPO2) levels for each person is visualized in the heatmap in **Fig 1D**. A majority (78%) experienced muscle or joint pain or tiredness, whereas only 26% had fever and 4% had breathing issues (**Supplemental Table 1**). None of them required hospital care, and at the second sampling time point after 14 days, all had a negative PCR test.

**Figure 1.**
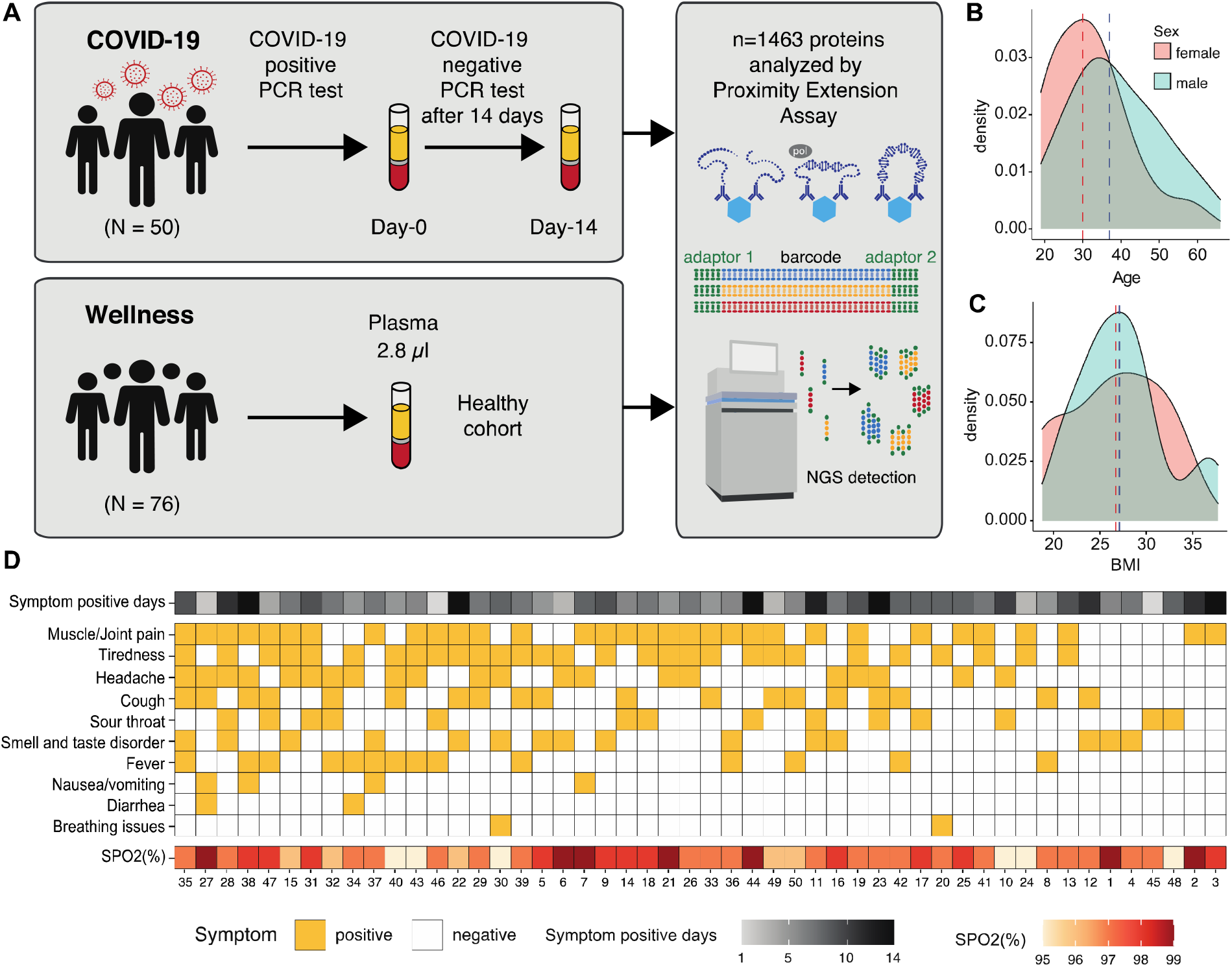
Overview of the study cohort. A. The study design with the sampling time points of the COVID-19 cohort as well as the wellness cohort from the S3WP program, which were both analyzed using the PEA-NGS method. B. The age distribution of the COVID-19 cohort. C. The BMI distribution of the COVID-19 cohort. D. Heatmap showing the symptoms of each individual of the COVID-19 cohort as well as the symptom positive days and the measured oxygen saturation (SPO2) levels in percentage (%).

### Next generation plasma proteome profiling

We used an approach for plasma protein profiling of the COVID-19 cohort where 1463 unique proteins were measured using the Olink platform (Olink® Explore 1536), which combines the PEA technology with Next Generation Sequencing (NGS) for read-out. The PEA-NGS technology allows for relative quantification of plasma protein expression levels which are calculated as Normalized Protein eXpression (NPX) values. A list with details about all analyzed plasma proteins is available in **Supplemental Table 2** and the complete table of NPX values for each protein is available in **Supplemental Table 3**. In **Fig 2A**, the expression profiles for each of the day-0 and day-14 samples based on all proteins were visualized using the dimensionality reduction method Uniform Manifold Approximation and Projection (UMAP) (McInnes, 2018). The resulting plot shows a separation between the two groups of samples, with most of the infection samples located together (red circle), but with some samples clustered at the individual level. The circular dendrogram in **Fig 2B** shows the result from hierarchical clustering of samples colored by sample group. Here, we see two smaller clusters with mainly day-0 or day-14 samples clustered together, respectively, which indicates similar protein signatures within each group. However, in most cases each individual is most closely clustered with itself, supporting the previous reports stating that each individual has a unique and stable global proteome profile (Tebani *et al*., 2020; Zhong *et al*., 2021). This is also evident in **Supplemental Fig 1A**, where the same dendrogram is colored by individual instead of sample group. A UMAP plot based on only day-0 samples shows that the global expression patterns cannot be explained by sex (**Supplemental Fig 1B**) or age (**Supplemental Fig 1C**) differences.

**Figure 2.**
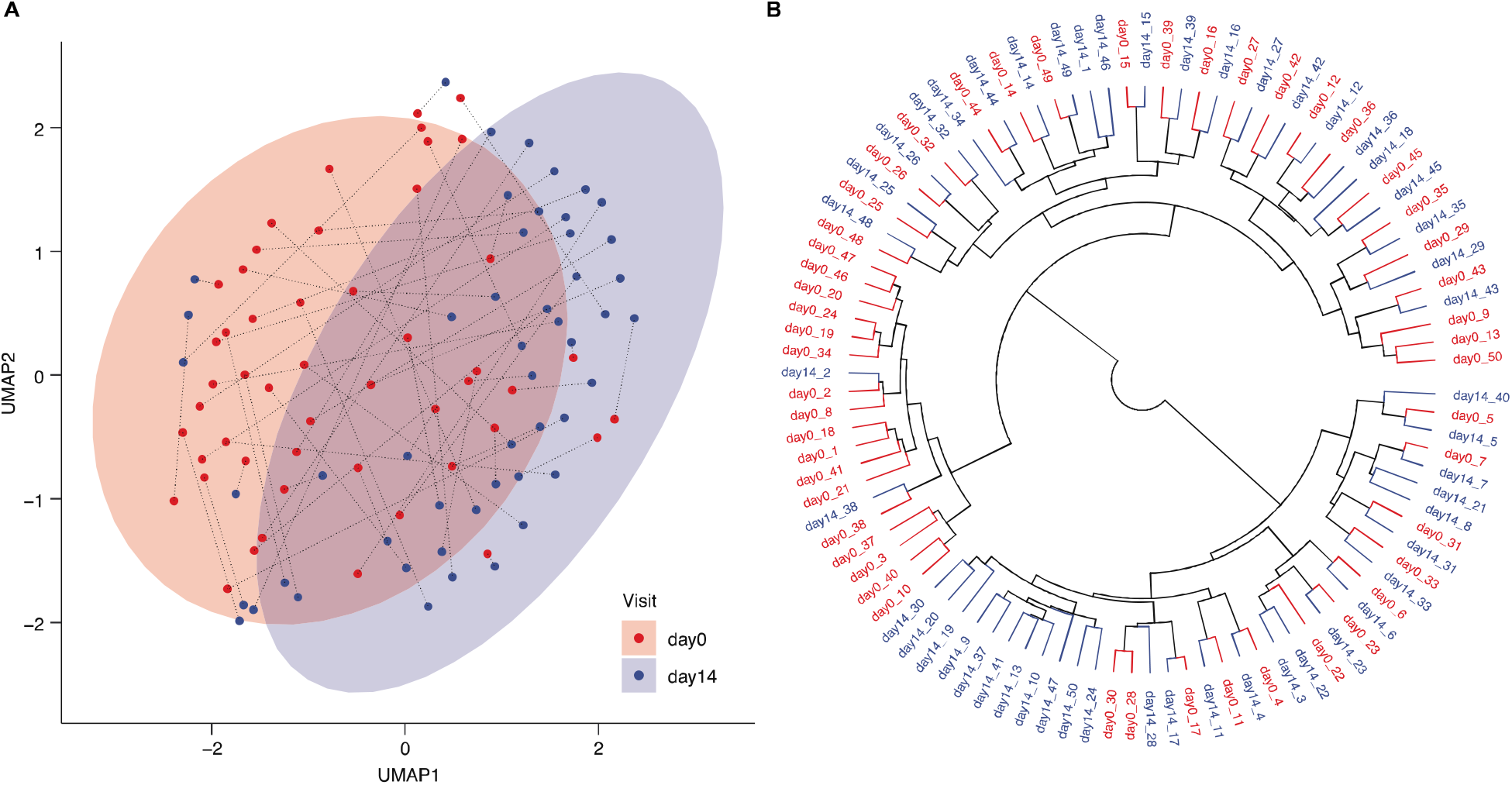
Clustering of COVID-19 samples. A. UMAP plot showing the distribution of day-0 and day-14 samples, with each individual connected by a dotted line. B. Dendrogram visualizing the results from hierarchical clustering of all samples. Day-0 samples are shown in red and day-14 samples in blue.

### Analysis of the plasma protein response to infection

We performed a multifactor analysis of variance (ANOVA) for all 1463 proteins to discover the most highly associated proteins to COVID-19, while also taking into consideration the effects of age, sex and BMI (**Fig 3A and Supplemental Table 4**). The most significantly associated protein with COVID-19 disease is scavenger receptor class B member 2 (SCARB2), which is a host cell receptor protein involved in virus entry and has recently been described in the context of SARS-CoV-2 (Patel *et al*., 2021). As expected from our previous studies (Tebani *et al*., 2020), the most highly associated protein with BMI is leptin (LEP) (**Fig 3A**). Cadherin related family member 2 (CDHR2) is the most significant sex-associated protein and is also associated with BMI (**Fig 3B**). Ectodysplasin A2 receptor (EDA2R) (**Fig 3C**) is most highly associated with age in our cohort supporting previous studies showing that this protein is linked to aging (Jeong *et al*, 2020; Ren & Kuan, 2020).

**Figure 3.**
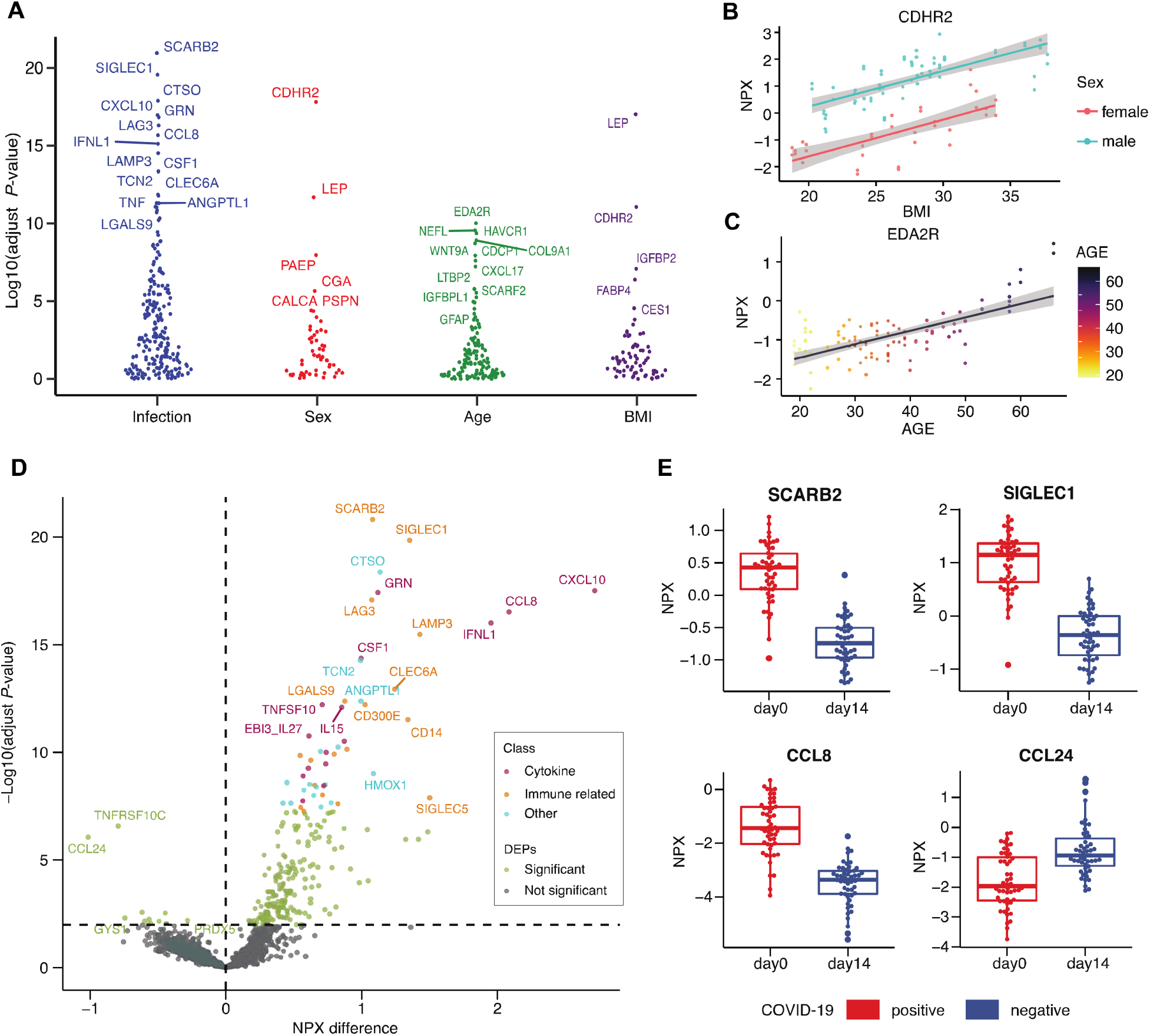
Proteins associated with COVID-19 infection. A. Results from multifactor ANOVA based on the factors COVID-19 infection (day-0 / day-14), age, sex and BMI, showing the most highly associated proteins with each factor. B. Example of a BMI-associated protein Leptin (LEP). C. Example of a sex-associated protein Cadherin related family member 2 (CDHR2). D. Volcano plot with differentially expressed proteins between day-0 and day-14 samples showing the difference in NPX values on the x-axis and -log10(adjusted p-value) on the y-axis. We show the manual annotation of the top 50 most significant proteins into groups of ‘cytokine’ (red), ‘immune related’ (orange) and ‘other’ (blue). E. Boxplots of examples of up-and down-regulated proteins in day-0 samples.

To further investigate which plasma proteins are most highly related to COVID-19 infection, we calculated the mean difference in expression and compared to the statistical significance based on the ANOVA results between the two groups of day-0 and day-14 samples for each protein. In the resulting volcano plot (**Fig 3D**), all proteins with adjusted p-value < 0.01 are considered significant (n=245) and the full list is provided in **Supplemental Table 4**. In addition, we performed a manual annotation of the biological function of the top 50 most significant proteins and classified them into three groups: (1) ‘cytokine’, (2) ‘immune related’, or (3) ‘other’ (**Table 1**). Interestingly, the Scavenger receptor class B member 2 (SCARB2) (**Fig 3E**), which is the most significant elevated plasma protein in the infected cohort, is reported as the cellular receptor for viral infection and responsible for viral entry (Chen *et al*, 2012). Among the proteins differentially expressed in the COVID-19 infection samples almost all are up-regulated during the infection, including many proteins related to cytokine response, for example Interferon lambda 1 (IFNL1) and the chemotactic factors C-X-C motif chemokine ligand 10 (CXCL10) and C-C motif chemokine ligand 8 (CCL8), which is known to play a role in neoplasia and inflammatory host responses (**Fig 3E**). Proteins related to other immune-related functions were also found, including Sialic acid binding Ig like lectin 1 (SIGLEC1), which functions as a macrophage-restricted adhesion molecule (**Fig 3E**), and Lymphocyte activating 3 (LAG3), which functions as an inhibitory receptor on antigen activated T-cells (Huard *et al*, 1994). Only two proteins are found to be significantly down-regulated during infections: (i) the C-C motif chemokine ligand 24 (CCL24), which is a cytokine involved in the inflammatory response (**Fig 3E**) and is a chemotactic for resting T-lymphocytes and eosinophils and (ii) the TNF receptor superfamily member 10c (TNFRSF10C), which is a receptor for the cytotoxic ligand TRAIL.

**Table 1.** Elevated plasma proteins in COVID-19 infected patients.

| Protein | UniProt description | Classification | NPX difference | adjust P-value |
| --- | --- | --- | --- | --- |
| SCARB2 | scavenger receptor class B member 2 | Immune related | 1.08 | 1.5E-21 |
| SIGLEC1 | sialic acid binding Ig like lectin 1 | Immune related | 1.35 | 1.4E-20 |
| CTSO | cathepsin O | Other | 1.14 | 4.2E-19 |
| CXCL10 | C-X-C motif chemokine ligand 10 | Cytokine | 2.72 | 3.1E-18 |
| GRN | granulin precursor | Cytokine | 1.12 | 3.8E-18 |
| LAG3 | lymphocyte activating 3 | Immune related | 1.08 | 8.3E-18 |
| CCL8 | C-C motif chemokine ligand 8 | Cytokine | 2.09 | 3.0E-17 |
| IFNL1 | interferon lambda 1 | Cytokine | 1.95 | 1.0E-16 |
| LAMP3 | lysosomal associated membrane protein 3 | Immune related | 1.43 | 3.3E-16 |
| CSF1 | colony stimulating factor 1 | Cytokine | 1.00 | 4.3E-15 |
| TCN2 | transcobalamin 2 | Other | 0.99 | 5.4E-15 |
| CLEC6A | C-type lectin domain containing 6A | Immune related | 1.24 | 1.2E-13 |
| ANGPTL1 | angiopoietin like 1 | Other | 0.99 | 4.2E-13 |
| LGALS9 | galectin 9 | Immune related | 0.88 | 4.2E-13 |
| CD300E | CD300e molecule | Immune related | 1.03 | 6.2E-13 |
| TNFSF10 | TNF superfamily member 10 | Cytokine | 0.71 | 6.2E-13 |
| IL15 | interleukin 15 | Cytokine | 0.85 | 8.1E-13 |
| CD14 | CD14 molecule | Immune related | 1.34 | 3.0E-12 |
| EBI3_IL2 | NA | Cytokine | 0.61 | 1.7E-11 |
| CX3CL1 | C-X3-C motif chemokine ligand 1 | Cytokine | 0.87 | 3.0E-11 |
| LGMN | legumain | Other | 0.83 | 5.7E-11 |
| CLEC4C | C-type lectin domain family 4 member C | Immune related | 0.89 | 7.1E-11 |
| TINAGL1 | tubulointerstitial nephritis antigen like 1 | Other | 0.70 | 9.1E-11 |
| CRLF1 | cytokine receptor like factor 1 | Cytokine | 0.74 | 1.0E-10 |
| PTX3 | pentraxin 3 | Immune related | 0.80 | 1.2E-10 |
| C1QA | complement C1q A chain | Immune related | 0.55 | 1.4E-10 |
| LILRA5 | leukocyte immunoglobulin like receptor A5 | Immune related | 0.62 | 2.3E-10 |
| IL18BP | interleukin 18 binding protein | Cytokine | 0.73 | 3.5E-10 |
| TNF | tumor necrosis factor | Cytokine | 0.61 | 5.4E-10 |
| HMOX1 | heme oxygenase 1 | Other | 1.09 | 9.9E-10 |
| IL18R1 | interleukin 18 receptor 1 | Cytokine | 0.57 | 1.3E-09 |
| ENTPD6 | ectonucleoside triphosphate diphosphohydrolase 6 (putative) | Other | 0.45 | 2.5E-09 |
| VWA1 | von Willebrand factor A domain containing 1 | Other | 0.62 | 3.1E-09 |
| ESM1 | endothelial cell specific molecule 1 | Other | 0.73 | 3.2E-09 |
| DLL1 | delta like canonical Notch ligand 1 | Immune related | 0.65 | 3.4E-09 |
| TNFSF13 | TNF superfamily member 13b | Cytokine | 0.72 | 3.6E-09 |
| FOLR2 | folate receptor beta | Other | 0.67 | 4.2E-09 |
| GAS6 | growth arrest specific 6 | Other | 0.58 | 5.8E-09 |
| LILRB4 | leukocyte immunoglobulin like receptor B4 | Immune related | 0.71 | 9.6E-09 |
| SEMA3F | semaphorin 3F | Other | 0.65 | 1.0E-08 |
| SIGLEC5 | sialic acid binding Ig like lectin 5 | Immune related | 1.50 | 1.3E-08 |
| TNFSF13 | TNF superfamily member 13 | Cytokine | 0.57 | 1.8E-08 |
| TPP1 | tripeptidyl peptidase 1 | Other | 0.77 | 2.2E-08 |
| ENTPD5 | ectonucleoside triphosphate diphosphohydrolase 5 | Other | 0.42 | 2.2E-08 |
| SMOC1 | SPARC related modular calcium binding 1 | Other | 0.48 | 2.2E-08 |
| BST2 | bone marrow stromal cell antigen 2 | Immune related | 0.82 | 2.5E-08 |
| FST | folistatin | Other | 0.70 | 3.5E-08 |
| VCAM1 | vascular cell adhesion molecule 1 | Immune related | 0.55 | 3.6E-08 |
| VSIG4 | V-set and immunoglobulin domain containing 4 | Immune related | 0.71 | 4.1E-08 |
| CD74 | CD74 molecule | Immune related | 0.58 | 5.4E-08 |

### Comparing mild/moderate COVID-19 disease profiles with severe disease

The expression levels of the 50 most significantly elevated proteins at COVID-19 infection (Table 1) are visualized as a heatmap in **Fig 4A**. As expected, most of the samples at infection (day 0) have a similar plasma protein profile (left part of the heatmap), while most of the plasma profiles after recovery (day 14) cluster together (right). However, there are samples with intermediate plasma proteins elevated at COVID-19 infection (middle). These identified proteins elevated at infection (day 0) were compared with the proteins elevated in patient with severe symptoms (Filbin *et al*., 2021) and the comparison (**Fig 4B)** shows that most proteins are elevated or down-regulated in a similar manner in patients with mild symptoms (this study) and severe symptoms (Filbin *et al*., 2021) (**Supplemental Table 5**). An example of an elevated plasma protein at infection is the chemokine CXCL10 which is involved in the stimulation of monocytes, natural killer and T-cell migration (**Fig 4C**). Similarly, an example (**Fig 4C**) of a protein down-regulated in patients both with mild and severe symptoms is the TNF receptor (TNFRSF10C), which is a receptor for the cytotoxic ligand TRAIL involved in the cellular apoptosis. The comparison thus suggests a good correlation in host plasma protein response in patients with mild and severe symptoms. However, there are some notably differences, in particular the SCARB2 protein mentioned above, which do not show elevated levels in the cohort from the patients with severe symptoms Filbin *et al*., 2021). There are also a group of proteins which are down regulated in our study who do not show down regulation in the patients with severe symptoms (**Fig 4B)**.

**Figure 4.**
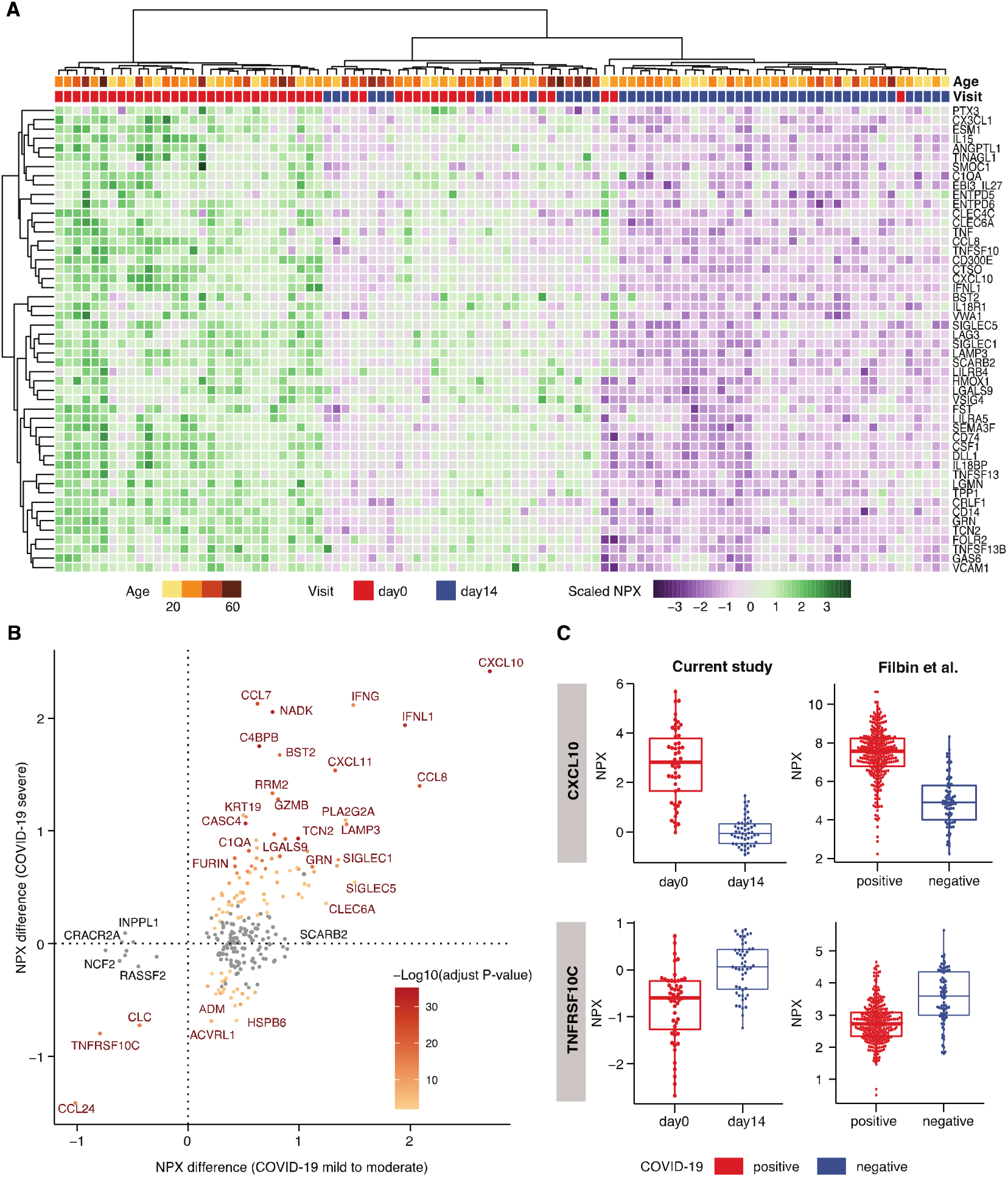
Analysis of the 50 most highly associated proteins to COVID-19 infections. A. Heatmap showing the expression levels of the 50 most significant proteins in all day-0 and day-14 samples, clustered based on expression in the 50 proteins. B. Scatterplot showing the difference in expression levels between day-0 and day-14 samples in our study on the x-axis, and the difference between COVID-19 positive and COVID-19 negative samples in the Filbin et al. (Filbin *et al*., 2021) study on the y-axis. All proteins with a significant difference in our study are shown (n=239). The color code depicts the -log10 adjusted p-value in the Filbin study, where the grey dots represent non-significant change. C. Boxplots of up- and down-regulated proteins in COVID positive and negative patients in both current and Filbin et al. studies.

### Comparison of the protein profiles between COVID-19 and healthy individuals

Next, a comparison with a healthy cohort of individuals analyzed with the same analytical platform as part of a wellness study (Zhong *et al*., 2021) was performed. In **Fig 5A**, the mean protein levels of the 50 most significant proteins (Table 1) in the infected patients (red) are shown as a radar plot and the levels are compared with the healthy individuals (green) and the same patients after recovery (blue). The results show the dramatic elevation of these proteins during acute infection, but also shows that in general these proteins have returned to healthy plasma levels after 14 days (recovery). As an example, the sialic acid binding Ig like lectin 1 (SIGLEC1/CD168) protein, which is found on circulating monocytes in COVID-19 (Doehn *et al*, 2021) and expression levels are associated with disease severity (**Fig 5B**). Next, a dimensionality reduction using UMAP was performed with the plasma profiles of the most significant proteins, including also the control group from the healthy population. The resulting UMAP plot (**Fig 5C**) shows distinct clusters of samples from the infected patients (red) and the healthy control group (green). As expected, most of the samples from the patients after recovery (blue) shows a pattern similar to the healthy control group, but interestingly some of the individuals have protein profiles similar to the infected patients. In **Fig 5D**, the same UMAP plot shows the individuals after the 14-day recovery, color coded according to age. Interestingly, the majority of the individuals with a “infected plasma profile” after 14 days are older suggesting a slower recovery in the older patients. In **Fig 5E**, the age distribution of the first group with plasma profiles resembling infected individuals are compared with the second group with individuals who have plasma profiles resembling the healthy control group. This demonstrates that there is an age-related difference in response to the COVID-19 infection since many of the older patients, despite that they have no symptoms, are not fully recovered after 14 days based on this exploration of their plasma proteins.

**Figure 5.**
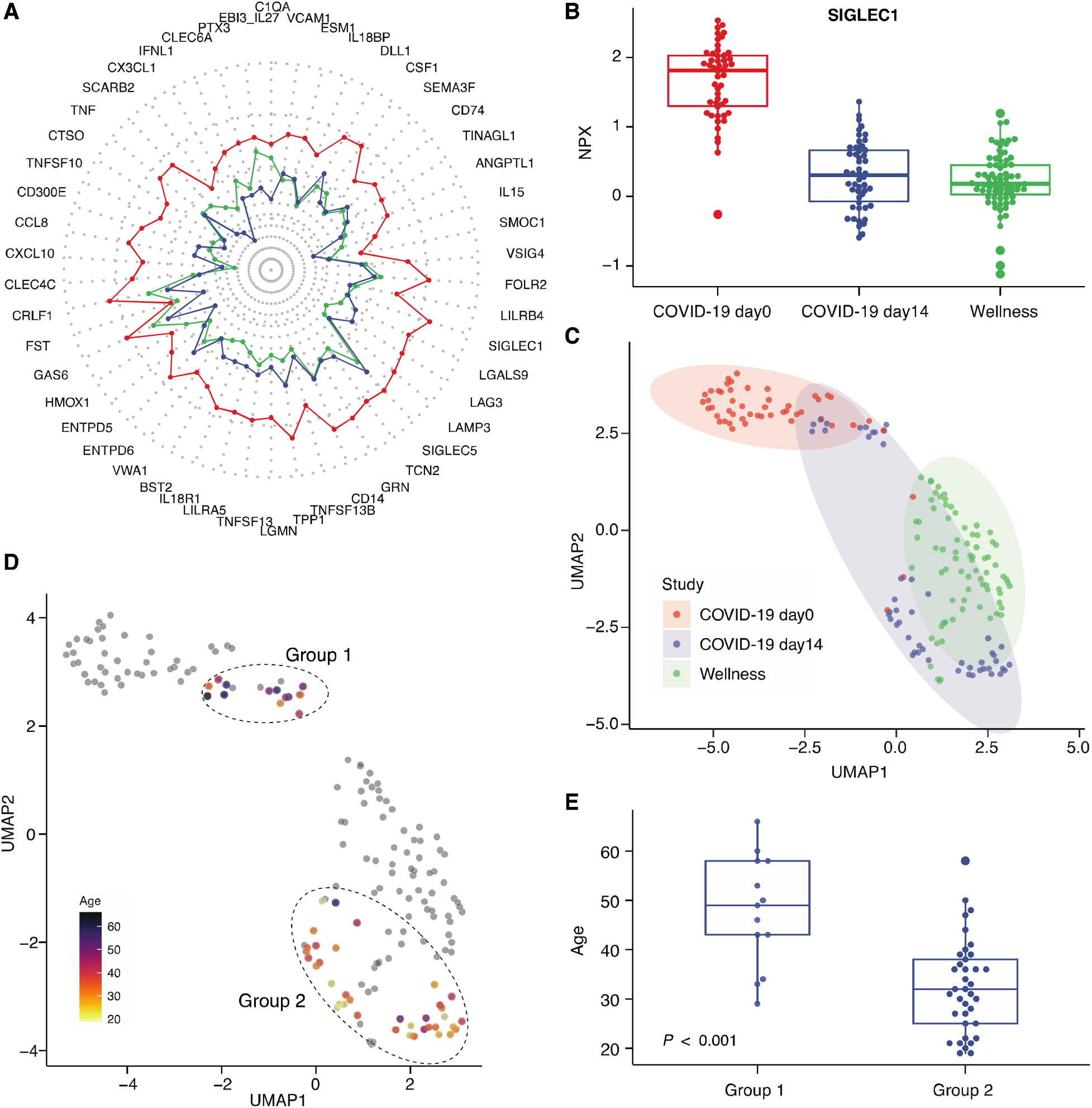
Analysis of the 50 most highly associated proteins to COVID-19 infections. A. Radar plot showing with the average expression levels of the 50 proteins for each of the three groups of samples (day-0, day-14 and wellness healthy cohort). B. Boxplot showing the expression levels of a differentially expressed protein sialic acid binding Ig like lectin 1 (SIGLEC1) in both the COVID-19 and wellness studies. C. UMAP plot showing the distribution of samples both from the COVID-19 cohort and the 76 samples from the wellness cohort. D. UMAP plot highlighting the age of the individuals in the two different groups of day-14 samples. E. Boxplot of the age distribution of the two different groups of day-14 samples and the p-value based on paired t-test.

## DISCUSSION

Here, we present a comprehensive overview of the host response during a COVID-19 infection based on proximity extension assay combined with next generation sequencing read-out, providing a sensitive and accurate multiplex analysis of plasma proteins. We have analyzed close to 1500 human proteins in non-hospitalized individuals with mild to moderate disease. The patients have been analysed after confirmation of infection with PCR-test and the plasma profile of each individual has been compared at the time of infection and after 14 days of recovery with no remaining symptoms of infection. The plasma levels were also compared to a healthy control cohort to further establish the response of plasma protein levels in these patients during recovery from the disease. More than 200 proteins were found to have significantly different plasma levels at the time of infection as compared to 14 days later. An analysis of the 50 most significant plasma proteins (Table 1) demonstrates that a majority of the proteins with different plasma levels at COVID-19 infection are cytokine- or immune-related. Interestingly, the analysis shows that many of the older patients retain a plasma profile similar to the acutely infected patients still after 14 days, despite having no symptoms of disease. The results suggest that there is an age-related difference in plasma profile recovery in the older patients.

We have also compared our results with the analysis of patients with severe symptoms described recently (Filbin *et al*., 2021; Patel *et al*., 2021). The comparison shows that a majority of the proteins show similar response to the infection independent of the severity of symptoms, demonstrating no difference in host response despite dramatic differences in symptoms. Thus, many immune related proteins are elevated at infection in both cases, such as chemochine ligand 10 (CXCL10), interferon gamma (IFNG), interferon lambda 1 (IFNL1) and chemokine ligand 8 (CCL8). However, there are differences in response for some proteins depending on the severity of symptoms of the corresponding patients. Most notably is the protein scavenger receptor class B member 2 (SCARB2) which in our study is the most significant elevated protein at infection, but is not shown to be elevated in the severe patients according to Filbin et al (Filbin *et al*., 2021), although the Patel study reported increasing levels of the protein in groups with severe or critical COVID-19 disease (Patel *et al*., 2021). SCARB2 is an interesting protein involved in membrane transportation and reorganization of endosomal and/or lysosomal compartments. This protein shows low tissue specificity (www.proteinatlas.org) (Uhlen *et al*, 2015) and studies have shown that the protein is involved in the pathogenesis of foot and mouth disease caused by enterovirus-71 and possibly by coxsackievirus A16. The question arises if this host cell receptor protein, involved in virus entry of enterovirus, could also be involved in SARS-CoV2 entry into the cell. The role in COVID-19 is completely unexplored and these results suggest that more in-depth studies should be performed to explore its involvement in COVID-19 infections.

In conclusion, we here describe a comprehensive plasma protein analysis of COVID-19 patients with mild to moderate symptoms. The analysis show that many proteins are elevated during COVID-19 infection and a comparison with earlier studies of patients with severe disease demonstrates similar plasma protein profiles independent of symptoms, but with some proteins differing in response. Interestingly, the analysis also reveals that older individuals have a slower recovery back to normal plasma levels after infection and the study demonstrates that many of these older patients display a “disease profile” even after 14 days of recovery, despite having no symptoms of disease. The study presented here demonstrates the usefulness of “next generation plasma protein profiling” to identify molecular signatures of importance for disease progression and to allow monitoring of disease during recovery from the infection. The results will facilitate further studies to understand the molecular mechanism of the host immune-related response of the SARS-CoV-2 virus.

## MATERIALS AND METHODS

### Participants

A total of 50 patients were randomly selected from a clinical trial cohort of 93 patients over 18 years of age, who had a PCR-confirmed COVID-19 test within the previous 24 h and were in stable condition not requiring hospitalization. Chest tomography was done to rule out pneumonia. Patients who had a partial oxygen saturation below 93% and required hospitalization after diagnosis were excluded. Treatment started on the day of diagnosis. All patients had a negative PCR test for COVID-19 on Day 14.

Participants for the randomized, open-label, placebo-controlled, phase-2 study for evaluating the efficacy and safety of combined metabolic activators in COVID-19 patients were recruited at the Umraniye Training and Research Hospital, University of Health Sciences, Istanbul, Turkey. Written informed consent was obtained from all participants before the initiation of any trial-related procedures. The safety of the participants and the risk–benefit analysis were overseen by an independent external data-monitoring committee. The trial was conducted in accordance with Good Clinical Practice guidelines and the principles of the Declaration of Helsinki. The study was approved by the ethics committee of Istanbul Medipol University, Istanbul, Turkey, and retrospectively registered at https://clinicaltrials.gov/ with Clinical Trial ID: NCT04573153. Patient information (patient number, date of birth, initials) was entered into the web-based randomization system, and the randomization codes were entered into the electronic case report form.

### Plasma protein profiling

Plasma proteins were analyzed using a multiplex Proximity Extension Assay (PEA) technology with high throughput sequencing readout (Olink Explore) (Assarsson *et al*., 2014; Zhong *et al*., 2020). As described before, the full library consists of specific antibodies targeting 1,472 proteins and 48 controls. Each antibody is labelled separately with unique PEA oligonucleotide probes, two separate and complementary sequences. The conjugated antibodies are mixed into four separate 384-plex panels (372 proteins and 12 internal controls used for QC and normalization) focused on inflammation, oncology, cardiometabolic and neurology proteins, respectively. The analytical performance of each of the protein assays included in the panel is carefully validated based on specificity, sensitivity, dynamic range, precision, scalability, endogenous interference and detectability (http://www.olink.com). Briefly, samples were randomized (different samples from the same individual were present within the same plate) and 2.8 µl of plasma were incubated overnight with antibodies conjugated to PEA probes at +4°C. Following the immune reaction, a combined extension and pre-amplification mix were added to the incubated samples at room temperature for PCR amplification. The PCR amplicons were thereafter pooled before a second PCR amplification step was performed with additions of individual sample index sequences. After pooling of samples, bead purification and QC of the generated libraries were followed on a Bioanalyzer. Finally, the sequencing was carried out using Illumina’s NovaSeq 6000 instrument using two S1 flow cells with 2 x 50 base read lengths. Counts of known barcode sequences were thereafter translated into normalized protein expression (NPX) units through a QC and normalization process. NPX is a relative protein quantification unit on a log2 scale and values are calculated from the number of matched counts on the NovaSeq run. Data generation of NPX consists of three main steps: normalization to the extension control (known standard), log2-transformation, and level adjustment using the plate control (plasma sample). Specifically engineered internal controls were added to each sample and are utilized to reduce intra-assay variability. These include one immuno-based control (incubation step) using a non-human assay, one extension control (extension step) composed of an antibody coupled to a unique DNA-pair always in proximity and, also, one amplification control (amplification step) based on a double stranded DNA amplicon. In addition, each sample plate includes sample controls used to estimate the precision (intra- and inter-CVs). Three negative controls (buffer only) are utilized to set background levels and calculate limit of detection (LOD), three plate controls (plasma pool) adjust levels between plates (thus improving inter-assay precision, allowing for optimal comparison of data derived from multiple runs), and finally two sample controls (reference plasma) are included to estimate precision. After quality control, a total of 1,459 proteins were included in the analysis.

### The wellness profiling study

The Swedish SciLifeLab SCAPIS Wellness Profiling (S3WP) program is based on the Swedish CArdioPulmonary bioImage Study (SCAPIS), which is a prospective observational study with 30,154 individuals enrolled at ages between 50 and 64 years from a random sampling of the general Swedish population16, from 2015 to 2018 (Tebani *et al*., 2020; Zhong *et al*., 2021; Zhong *et al*., 2020). In total, 101 healthy individuals were recruited. Extensive phenotype characterization of the subjects was conducted before the study to establish the inclusion and exclusion criteria for the definition of ‘healthy’ subjects. The exclusion criteria in the S3WP program included: 1) previously received health care for myocardial infarction, stroke, peripheral artery disease or diabetes, 2) presence of any clinically significant disease which, in the opinion of the investigator, may interfere with the results or the subject’
ss ability to participate in the study, 3) any major surgical procedure or trauma within 4 weeks of the first study visit, or 4) medication for hypertension or hyperlipidemia. The study is approved by the Ethical Review Board of Göteborg, Sweden (registration number 407-15). All participants provided written informed consent. The study protocol conforms to the ethical guidelines of the 1975 Declaration of Helsinki. As described before, a total of 76 subjects were randomly selected from the wellness study to investigate the plasma levels of proteins using PEA-NGS (Olink Explore) technology (Zhong *et al*., 2021).

### Normalization of the plasma proteome profiling data

To allow for comparison of the two cohorts, the protein expression profiles from the wellness study were normalized to the current study using intensity normalization based on control samples (n = 20, wellness study; n = 8, COVID-19 study) (see details in http://www.olink.com). In brief, 1) for each study and assay, the study specific median value was calculated based on all control samples; 2) for each assay, an assay-specific normalization factor was estimated by calculating the median level of the pairwise differences for each of the control samples; 3) for each assay in the wellness study, the assay-specific normalization factor was added to the original NPX value, to normalize it to the current study.

### Statistical Analysis

All data analysis and visualization was performed using R (v3.6.3)(R Development Core Team, 2021). Differential expression analysis was carried out using analysis of variance (ANOVA) method with the built-in R function “anova()”. False discovery rate (FDR) was calculated by using p.adjust() function in R, which uses Benjamini−Hochberg method. Proteins with FDR < 0.01 were considered as differentially expressed proteins. Uniform Manifold Approximation and Projection (UMAP)(McInnes, 2018) was performed based on scaled NPX values using the R packages umap (Konopka, 2020). The hierarchical clustering result visualized in dendrograms was based on Pearson correlation and created by first calculating a correlation matrix of Pearson’s ρ between all analyzed samples. The correlation was converted to a distance metric (1 – ρ) and was clustered using the Ward2 algorithm. Circular dendrogram and radar chart were generated using R packages circlize (Gu *et al*, 2014) and fsmb (Nakazawa & Nakazawa, 2019).

## Data Availability

The COVID-19 cohort plasma profiling dataset and metadata are available in the supplementary material. The S3WP healthy cohort dataset has been deposited with the Swedish National Data Service (www.snd.se, a data repository certified by Core Trust Seal): doi: 10.5878/rdys-mz27. The dataset can be made available for validation purposes by contacting. Data access will be evaluated according to Swedish legislation. Data access for research related questions in the S3WP program can be made available by contacting the corresponding author.

## ACKNOWLEDGMENTS

This work was financially supported by Knut and Alice Wallenberg Foundation.

## SUPPLEMENTARY MATERIAL

The supplementary material consists of five Supplemental Tables and two Supplemental Figures.

## AUTHOR CONTRIBUTIONS

MU, WZ and LF conceived and designed the analysis. AM, MA, FE and OA collected and contributed data to the study. WZ, LF and MU performed the data analysis. MU, WZ and LF drafted the manuscript. All authors read and approved the final manuscript.

## REFERENCES

Assarsson E, Lundberg M, Holmquist G, Bjorkesten J, Thorsen SB, Ekman D, Eriksson A, Rennel Dickens E, Ohlsson S, Edfeldt G et al (2014) Homogenous 96-plex PEA immunoassay exhibiting high sensitivity, specificity, and excellent scalability. PLoS One 9: e95192

Chen P, Song Z, Qi Y, Feng X, Xu N, Sun Y, Wu X, Yao X, Mao Q, Li X et al (2012) Molecular determinants of enterovirus 71 viral entry: cleft around GLN-172 on VP1 protein interacts with variable region on scavenge receptor B 2. J Biol Chem 287: 6406–6420

Del Valle DM, Kim-Schulze S, Huang HH, Beckmann ND, Nirenberg S, Wang B, Lavin Y, Swartz TH, Madduri D, Stock A et al (2020) An inflammatory cytokine signature predicts COVID-19 severity and survival. Nat Med 26: 1636–1643

Doehn JM, Tabeling C, Biesen R, Saccomanno J, Madlung E, Pappe E, Gabriel F, Kurth F, Meisel C, Corman VM et al (2021) CD169/SIGLEC1 is expressed on circulating monocytes in COVID-19 and expression levels are associated with disease severity. Infection

Filbin MR, Mehta A, Schneider AM, Kays KR, Guess JR, Gentili M, Fenyves BG, Charland NC, Gonye ALK, Gushterova I et al (2021) Longitudinal proteomic analysis of plasma from patients with severe COVID-19 reveal patient survival-associated signatures, tissue-specific cell death, and cell-cell interactions. Cell Rep Med: 100287

Gu Z, Gu L, Eils R, Schlesner M, Brors B (2014) circlize Implements and enhances circular visualization in R. Bioinformatics 30: 2811–2812

Gummesson A, Bjornson E, Fagerberg L, Zhong W, Tebani A, Edfors F, Schmidt C, Lundqvist A, Adiels M, Backhed F et al (2021) Longitudinal plasma protein profiling of newly diagnosed type 2 diabetes. EBioMedicine 63: 103147

Gupta A, Madhavan MV, Sehgal K, Nair N, Mahajan S, Sehrawat TS, Bikdeli B, Ahluwalia N, Ausiello JC, Wan EY et al (2020) Extrapulmonary manifestations of COVID-19. Nat Med 26: 1017–1032

Hou X, Zhang X, Wu X, Lu M, Wang D, Xu M, Wang H, Liang T, Dai J, Duan H et al (2020) Serum Protein Profiling Reveals a Landscape of Inflammation and Immune Signaling in Early-stage COVID-19 Infection. Mol Cell Proteomics 19: 1749–1759

Huang C, Wang Y, Li X, Ren L, Zhao J, Hu Y, Zhang L, Fan G, Xu J, Gu X et al (2020) Clinical features of patients infected with 2019 novel coronavirus in Wuhan, China. Lancet 395: 497–506

Huard B, Tournier M, Hercend T, Triebel F, Faure F (1994) Lymphocyte-activation gene 3/major histocompatibility complex class II interaction modulates the antigenic response of CD4+ T lymphocytes. Eur J Immunol 24: 3216–3221

Jeong I, Lim JH, Park JS, Oh YM (2020) Aging-related changes in the gene expression profile of human lungs. Aging (Albany NY) 12: 21391–21403

Konopka T (2020) umap: Uniform Manifold Approximation and Projection.

Lucas C, Wong P, Klein J, Castro TBR, Silva J, Sundaram M, Ellingson MK, Mao T, Oh JE, Israelow B et al (2020) Longitudinal analyses reveal immunological misfiring in severe COVID-19. Nature 584: 463–469

Mathew D, Giles JR, Baxter AE, Oldridge DA, Greenplate AR, Wu JE, Alanio C, Kuri-Cervantes L, Pampena MB, D’Andrea K et al (2020) Deep immune profiling of COVID-19 patients reveals distinct immunotypes with therapeutic implications. Science 369

McInnes L, Healy, J. & Melville, J, 2018. UMAP: Uniform Manifold Approximation and Projection for Dimension Reduction

Mehta P, McAuley DF, Brown M, Sanchez E, Tattersall RS, Manson JJ, Hlh Across Speciality Collaboration UK (2020) COVID-19: consider cytokine storm syndromes and immunosuppression. Lancet 395: 1033–1034

Nakazawa M, Nakazawa MM (2019) Package ‘fmsb’. Retrived from https://cranr-projectorg/web/packages/fmsb/fmsbpdf

Patel H, Ashton NJ, Dobson RJB, Andersson LM, Yilmaz A, Blennow K, Gisslen M, Zetterberg H (2021) Proteomic blood profiling in mild, severe and critical COVID-19 patients. Sci Rep 11: 6357

R Development Core Team, 2021. R: A Language and Environment for Statistical Computing. R Foundation for Statistical Computing.

Ren X, Kuan PF (2020) RNAAgeCalc: A multi-tissue transcriptional age calculator. PLoS One 15: e0237006

Tebani A, Gummesson A, Zhong W, Koistinen IS, Lakshmikanth T, Olsson LM, Boulund F, Neiman M, Stenlund H, Hellstrom C et al (2020) Integration of molecular profiles in a longitudinal wellness profiling cohort. Nat Commun 11: 4487

Uhlen M, Fagerberg L, Hallstrom BM, Lindskog C, Oksvold P, Mardinoglu A, Sivertsson A, Kampf C, Sjostedt E, Asplund A et al (2015) Proteomics. Tissue-based map of the human proteome. Science 347: 1260419

Williamson EJ, Walker AJ, Bhaskaran K, Bacon S, Bates C, Morton CE, Curtis HJ, Mehrkar A, Evans D, Inglesby P et al (2020) Factors associated with COVID-19-related death using OpenSAFELY. Nature 584: 430–436

Wu Z, McGoogan JM (2020) Characteristics of and Important Lessons From the Coronavirus Disease 2019 (COVID-19) Outbreak in China: Summary of a Report of 72314 Cases From the Chinese Center for Disease Control and Prevention. JAMA 323: 1239–1242

Zhong W, Edfors F, Gummesson A, Bergstrom G, Fagerberg L, Uhlen M (2021) Next generation plasma proteome profiling to monitor health and disease. Nat Commun 12: 2493

Zhong W, Gummesson A, Tebani A, Karlsson MJ, Hong MG, Schwenk JM, Edfors F, Bergstrom G, Fagerberg L, Uhlen M (2020) Whole-genome sequence association analysis of blood proteins in a longitudinal wellness cohort. Genome Med 12: 53

