## Supplementary Material for "Next generation plasma proteome profiling of COVID-19 patients with mild to moderate symptoms"

**The PDF file includes:**

Supplementary Figure 1. Clustering of COVID-19 patients..

Supplementary Figure 2. Correlation between the 50 most significantly elevated proteins at COVID-19 infection

**Other Supplementary Material for this manuscript includes the following:**

Supplementary Table 1. Description of the COVID-19 patients

Supplementary Table 2. Full list of the analyzed plasma proteins

Supplementary Table 3. Plasma proteome profiling of the COVID-19 patients

Supplementary Table 4. Full list of the ANOVA results

Supplementary Table 5. Comparison of the differentially expressed proteins in mild-to-moderate and severe COVID-19 patients

**Supplemental Figure 1.**

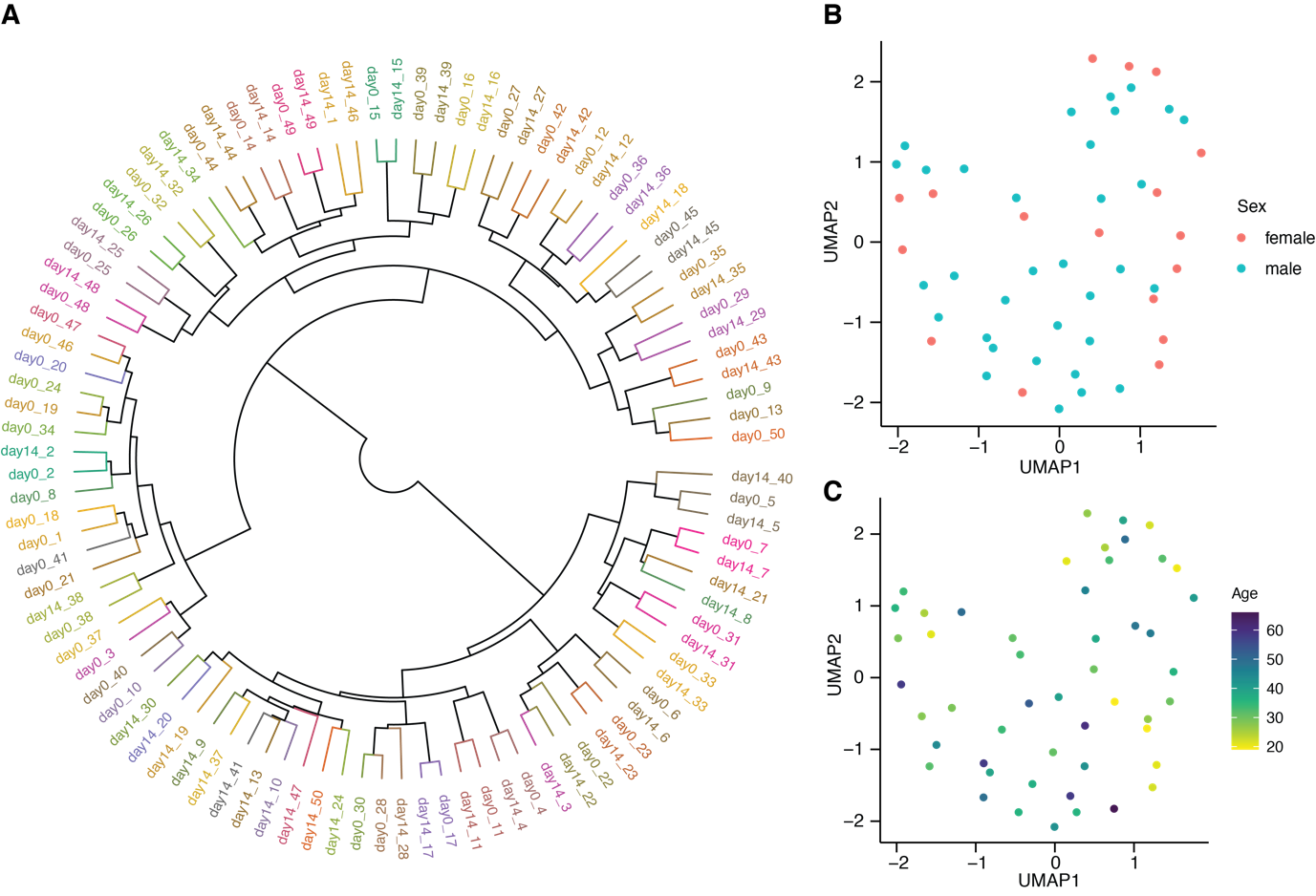

**Supplemental Figure 1. Clustering of COVID-19 patients.**

A. Dendrogram visualizing the results from hierarchical clustering of all samples. The color code indicates individuals.

B. UMAP plot showing the distribution of day-0 samples with ongoing COVID-19 infection. The color code indicates females and males.

C. UMAP plot showing the distribution of day-0 samples with ongoing COVID-19 infection. The color code indicates age differences.

**Supplemental Figure 2**
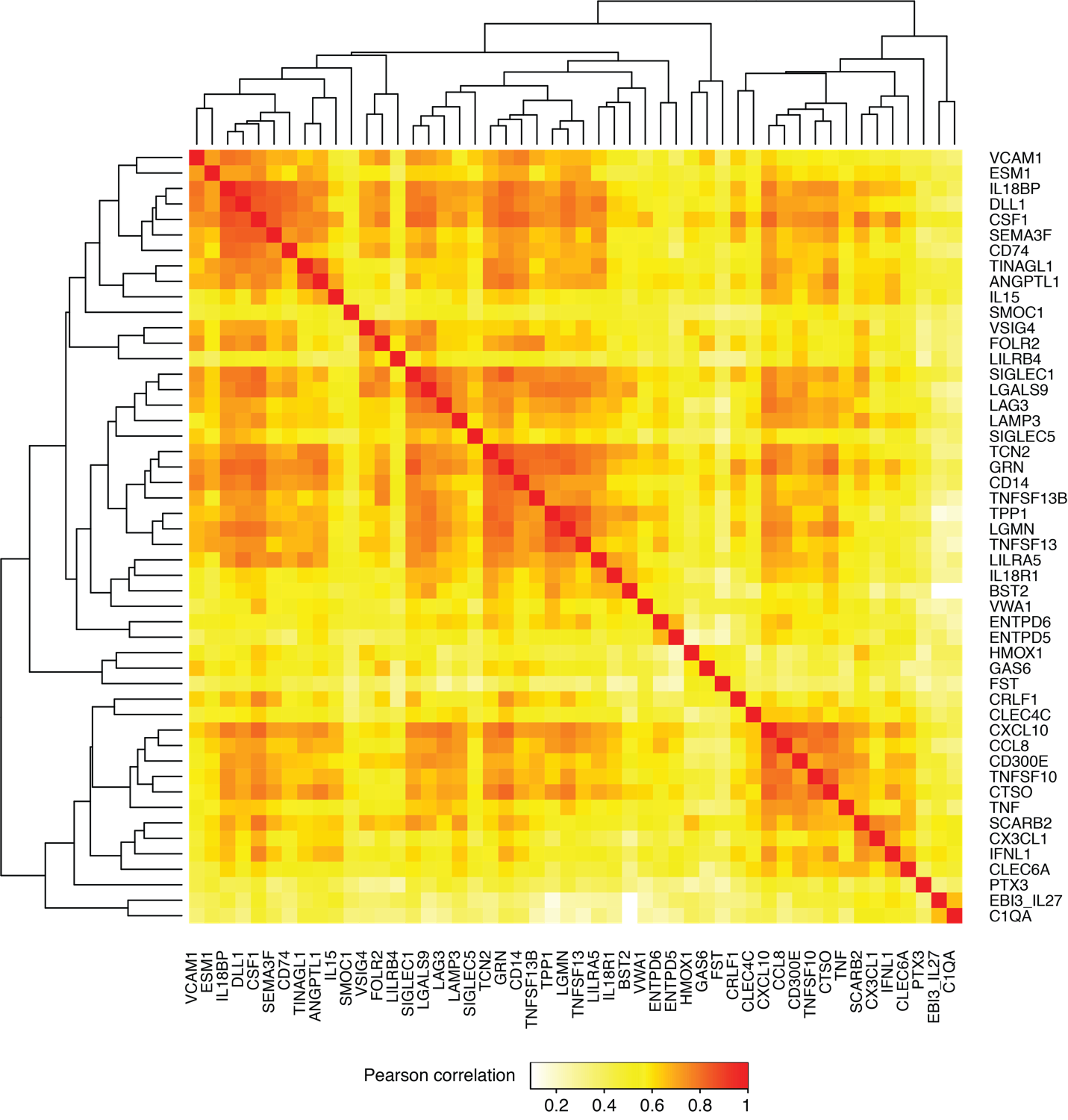

**Supplemental Figure 2. Correlation between the 50 most significantly elevated proteins at COVID-19 infection.**

Heatmap showing pairwise Pearson correlation between the expression profiles for the 50 proteins analyzed. The dendrogram is clustered by the correlation distance using the Ward2 algorithm.
